# User Perceptions of Individually-Tailored Health Information in Digital Apps: Development of a Scale

**DOI:** 10.1101/2025.10.25.25338794

**Authors:** Raymond L. Ownby, Rosemary Davenport, Joshua Caballero

## Abstract

**Objectives:** Individually tailored health information is thought to have greater effects on patient behavior than generic advice because it is more personally relevant. Most digital health studies, however, do not actually measure the effect of tailoring on study outcomes. To address this gap, we created the Success in Tailoring (SIT) scale which assesses how users perceive information as relevant, useful, and actionable.

**Methods:** The SIT items were chosen to reflect theoretical work on relevance and elements of the Elaboration Likelihood Model. It was administered to participants in a study of a mobile app providing tailored information about chronic disease self-management to persons 40 years of age and older with low health literacy. Participants responded immediately after completing the study intervention and again three months later. Psychometric analyses focused on the measure’s reliability, factor structure, and convergent and divergent validity other measures thought to be related and unrelated to it. We assessed test-retest reliability and factorial invariance over administration, and whether the measure predicted changes in key study outcomes.

**Results:** Analyses were based on responses from 275 participants. The SIT’s internal consistency was good, and test-retest reliability was acceptable. Exploratory factor analysis suggested a single-factor solution, although subsequent confirmatory analyses revealed that a bifactor solution with a robust general factor and two minor subfactors fit the data best. The scale was significantly correlated with measures related to its underlying concept and unrelated to measures not related to it, such as physical and cognitive status. Configural and metric, but not scalar, factorial invariance, were confirmed. SIT scores were related to change in activation and disease-management self-efficacy over the course of the study. Confirmatory bifactor analyses supported treating the SIT as essentially unidimensional, with a single total score providing a reliable and valid index of perceived tailoring.

**Conclusion:** The SIT gives researchers a straightforward means of capturing whether participants feel information is successfully tailored to them. It may be helpful in explaining how personalization, a key feature of many digital health apps, may be related to outcomes.

## 1 Introduction

Tailored health communication has long been recognized as more effective than generic information in promoting patient engagement and behavior change. Health communications that take account of a person’s own interests and needs are perceived as more relevant, processed more deeply, and are more likely to produce behavior change. In the context of digital health interventions, tailoring is particularly important, as apps and online tools can adapt content dynamically to user responses and behaviors.

While extensive research supports the usefulness of tailoring [1, 2], few studies have quantified it when implemented. Because of this, we cannot tell whether users experience the content as personal and if tailoring drives observed effects. Research on digital tools often still points to clicks, logins, or general outcomes and treats them as evidence of effectiveness [3]. To address this issue, we created the Success in Tailoring (SIT) scale, a brief measure of how well participants feel intervention content fits their needs and circumstances. The scale grew out of the Elaboration Likelihood Model (ELM) and was informed by qualitative work with adults managing multiple chronic conditions [4]. Items were chosen to reflect an underlying model of the dimensions of information relevance [5, 6], including the extent to which a person viewed the tailored information as related to their situation, their needs, and the extent to which it was useful.

The SIT was used in a study of a tailored information app for chronic disease self-management [7], and we showed, for example, that users’ SIT scores were related to change in stress, which was in turn related to improved quality of life [8]. The SIT has promise, but it is still missing a more extensive formal validation. Testing its reliability, factor structure, and links to related measures like patient activation, self-efficacy, and health-related quality of life would give the scale a stronger foundation. The purpose of this paper is to report this more extensive analysis of the reliability and validity of the SIT.

## 2 Methods

### 2.1 Parent study

#### 2.1.1 Participants and Procedures

In the study for which the SIT was developed, participants were adults aged 40 and older with at least one chronic health condition and low health literacy. They were recruited from local clinics, previous studies, and community outreach [7]. Eligibility required less than a college education and a score below the 8th-grade cutoff on the Rapid Estimate of Adult Literacy in Medicine or REALM [9]. Participants represented a wide range of chronic conditions, including both physical and mental health diagnoses, and all were receiving treatment with at least one prescribed or recommended medication.

The parent study evaluated a chronic disease self-management (CDSM) app designed for individuals with low health literacy. The app featured three versions, each presenting the health content at different reading levels (8th, 6th, and 3rd grade, the latter with audio narration). Content was developed by a multidisciplinary team and included extensive qualitative interviewing of relevant stakeholders [4] and was refined through multiple rounds of usability testing with the target population. Participants completed an extensive baseline assessment—including demographic information, medical and educational history, and standardized measures of reading ability and health literacy—either individually-administered or via audio computer-assisted self-interview software to minimize literacy-related response bias.

Health information related to chronic disease self-management of common problems related to multimorbidity was provided in a series of modules focused on topics such as treatment adherence, sleep, mood, fatigue, and pain. Each module consisted of a series of screens providing information on the topic. Individual tailoring was accomplished at the beginning of each module by asking participants to respond to questions related to difficulties in each area. For example, in the sleep module participants completed questions that asked about problems falling or staying asleep or waking up too early. They also responded to queries about how refreshing their sleep was and how sleep problems affected their daily activities. Later in the module, participants were provided with information related to the difficulties they reported, such as relaxation techniques or mediation for difficulties falling asleep.

For outcome assessment, widely used and validated instruments were administered at baseline, immediately following the intervention, and three months post-intervention. These included measures of patient activation [10], self-efficacy [11], health-related quality of life [12], and medication adherence [13]. The measure under validation in the present study was embedded within this comprehensive assessment protocol. A total of 275 participants responded to the SIT immediately after completing the study intervention, and 240 completed it at the three-month follow-up.

#### 2.1.2 Item Development

We developed the SIT items with the idea that people use different cues when they decide whether health information feels important. Saracevic’s reviews of relevance were our starting point, and we also kept the Elaboration Likelihood Model in mind as a way of thinking about personal relevance. See Table 1 for links between items, dimensions of relevance, and the ELM.

**Table 1.** Success in Tailoring (SIT) items mapped to relevance frameworks and the Elaboration Likelihood Model.

| SIT Item | Manifestation(s) of Relevance | Clues/Criteria | Stratified Model Layer | ELM Link |
| --- | --- | --- | --- | --- |
| I could relate to the information. | Situational, socio-cognitive | Use/situational match; cognitive match | Cognitive, affective, situational | Personal relevance → central route processing |
| The information was about people like me. | Socio-cognitive | Use/situational; belief match (identity/credence) | Socio-cognitive | Self-relevance cue; identification with message |
| The information would be useful to someone like me. | Situational | Use/situational match; topical | Situational | Relevance to personal goals enhances elaboration |
| The information was about the kind of problems people like me have. | Topical + socio-cognitive | Content; use/situational | Cognitive + situational | Topic + identity match increases elaboration likelihood |
| The information was easy to understand. | Cognitive | Cognitive match (understanding/effort) | Cognitive | Comprehension facilitates elaboration |
| The information would work for me. | Situational/utility | Use/situational (utility) | Situational | Perceived applicability strengthens motivation to act |
| The information gave me something to think about. | Cognitive + affective | Cognitive match (novelty); affective match (engagement) | Cognitive + affective | Message elaboration (deep thought) |
| I want to try out some of the things talked about. | Affective/motivational | Affective match; use/situational (intended use) | Affective, situational | Intention → behavior (ELM outcome of central route) |

Some of the items are clearly direct, like asking whether the material was “about people like me” or “about the kind of problems people like me have.” That was meant to tap the social and topical aspect of health information—whether people see themselves reflected in the content. Others focus more on the practical side, such as whether the information seemed useful or whether it “would work” for them. We also added an item about whether it was “easy to understand,” a basic check on comprehension or cognitive fit. Finally, several items go further, for instance, “gave me something to think about” or “I want to try out some of the things.” Those are intended to assess a participant’s motivation and affect—did the material generate interest or a plan to act?

Overall, then, the items assess the main aspects of relevance—cognitive, practical, and affective— with the kinds of clues people usually use to judge information. In relation to the Elaboration Likelihood Model, the items follow its elements. Starting with personal relevance and comprehension, moving through elaboration (“gave me something to think about”), and ending with intention (“I want to try out…”). The scale is therefore tied to both relevance theory and mechanisms thought to connect personalization with changes in health behavior.

### 2.2 Psychometric Evaluation

#### 2.2.1 Measures

##### Success in Tailoring (SIT)

The SIT was designed as a brief measure of participants’ perceptions of health information as personally relevant. Participants were asked to rate each item on a five-point Likert-type scale, and responses were summed to create a total score. Higher scores indicate greater perceived individual tailoring of the information.

###### External measures

To evaluate validity, we included several established constructs. The Patient Activation Measure (PAM) [10] and the Chronic Disease Self-Efficacy Scale (CDSE) [11] were used as convergent measures, as both assess constructs expected to relate closely to tailoring perceptions. Divergent validity was tested using scales less directly tied to tailoring, such as cognitive and academic measures and physical assessments. Predictive validity was assessed by evaluating the SIT’s ability to predict change in outcome measures over time.

#### 2.2.2 Procedure

The SIT was administered at two points: immediately after participants completed the intervention and again three months later. Cognitive and academic measures were administered only at baseline, and physical status (height, weight, blood pressure, walking speed) was also only assessed at baseline. Ethical approval for the study was obtained from the Institutional Review Boards of Nova Southeastern and Emory Universities. Participants gave verbal consent for screening, and all participants gave written informed consent for further participation.

#### 2.2.3 Analytic Strategy

Descriptive statistics were computed in SPSS version 29. Most other analyses were completed using packages in R (version 4.3.1). Mplus version 9 was used to create latent growth curve models. To evaluate the dimensionality of the SIT, we estimated a confirmatory bifactor model in which all items loaded on a general perceived-tailoring factor as well as on one of several content-specific group factors representing different aspects of tailored communication. Factors were specified as orthogonal. Model fit was compared with one- and two-factor solutions using robust maximum likelihood estimation (MLR) in *lavaan* [14]. To assess the relative strength of the general versus group factors, we calculated bifactor indices including the Explained Common Variance (ECV), Percent Uncontaminated Correlations (PUC), Omega Total (ΩT), Omega Hierarchical for the general (ΩH and group factors (ΩHS), construct replicability (H), and factor determinacy (FD) using the *BifactorIndicesCalculator* [15] and *semTools* [16] packages in R. Thresholds recommended in the literature (e.g., ECV ≥ 0.60, PUC ≥ 0.70, ΩH ≥ 0.70) were used to evaluate whether the measure could be considered essentially unidimensional [17, 18]. Together, these indices provided a comprehensive picture of the SIT’s dimensional structure and the degree to which a single score could represent the underlying construct of perceived tailoring.

##### 2.2.3.1 Exploratory Factor Analysis (EFA)

An exploratory factor analysis was used to examine the structure of the SIT items. Analyses relied on the *psych* [19] and *nFactors* [20] packages. The number of factors was guided by parallel analysis [21]. Principal axis factoring was used with evaluation of both varimax and oblimin rotations.

##### 2.2.3.2 Confirmatory Factor Analysis (CFA)

Competing models (one-factor, two-factor, and bifactor) were tested in *lavaan* [14]. Fit was evaluated using multiple indices: the Comparative Fit Index (CFI), Tucker–Lewis Index (TLI), Root Mean Square Error of Approximation (RMSEA) with 90% confidence interval, and the Standardized Root Mean Square Residual (SRMR). Because item distributions were moderately skewed, all latent variable analyses were conducted using robust maximum likelihood estimation (MLR), which provides standard errors and χ² statistics robust to non-normality.

##### 2.2.3.3 Longitudinal Measurement Invariance

Measurement invariance across the two administrations was evaluated in *lavaan*. Models were estimated using robust maximum likelihood (MLR) in *lavaan*, which provides standard errors and fit indices corrected for non-normality (Satorra–Bentler scaled) [22]. Configural, metric, and scalar models were estimated, and changes in fit indices were used as criteria for invariance (Δ CFI ≤ .01, Δ RMSEA ≤ .015 [23, 24]) as well as change in the likelihood ratio chi-square. Robust chi-square difference tests (via *lavTestLRT* from the *lavaan* package) were used to compare nested models, using the Satorra–Bentler correction for MLR estimation.

##### 2.2.3.4 Reliability Analyses

Test–retest reliability was estimated with intraclass correlations (ICCs) using reliability analysis in SPSS 29. Both absolute agreement [ICC(A,1)] and consistency [ICC(C,1)] were reported with 95% confidence intervals. Precision was further described with the standard error of measurement (SEM) and the smallest detectable change (SDC) at the 95% level. Agreement at the individual level was assessed with Bland–Altman plots [25] produced in *ggplot2* [26], which showed mean bias, 95% limits of agreement, and a regression line to test for proportional bias. We also produced a spaghetti plot of SIT scores at the two time points to visually assess change over time.

##### 2.2.3.5 Convergent and Divergent Validity

Convergent validity was examined by correlating SIT scores with constructs expected to be related, including perceived usefulness of the app (Usefulness scale of a Technology Acceptance Model-based user feedback scale) [27]; patient activation (Patient Activation Measure, PAM)[10]; participant level of health literacy (Test of Functional Health Literacy in Adults, TOFHLA, and FLIGHT/VIDAS Health Literacy Measure, FVA) [28, 29]; internal locus of control (Internal Control subscale of the Multidimensional Health Locus of Control Scale, MHLC) [30]; disease management self-efficacy (Chronic Disease Self-Efficacy Scale, CDSE) [11]. Divergent validity was examined as correlations of constructs expected to be unrelated, including participant self-report of annual income; stress (Perceived Stress Scale, PSS) [31]; depressive symptoms (Center for Epidemiological Studies—Depression scale, CESD) [32]; participant report of total number of chronic health conditions; age in years; participant systolic blood pressure before completing the 10-meter walk test and time taken in the 10-meter walk test [33]. Pearson correlations were computed with base R functions.

##### 2.2.3.6 Predictive Validity

Predictive validity was tested using latent growth models in Mplus 9 [34]. Latent growth curve models, including the parent study’s primary outcomes of patient activation and disease management self-efficacy, were constructed in several stages. In the first stage, growth curves were calculated to assess whether the variables had changed over the course of the study by testing whether the slope was significantly different from zero. For each growth curve, its intercept and slope were then regressed on potentially important covariates, age, gender, and race to account for potential confounding variables. Finally, the slopes were regressed on the SIT score to determine whether SIT scores were related to change in outcomes over the course of the study. Health-related quality of life at the three-month follow-up (QOL; SF36 General Health scale) [12] was then regressed on the slopes, and the indirect effect of the SIT on QOL was evaluated.

## 3 Results

### 3.1 Sample Characteristics

Descriptive information for the sample is presented in Table 2. The sample included 275 participants aged 40 years and older with low health literacy and at least one chronic health condition for which they were being treated.

**Table 2.**
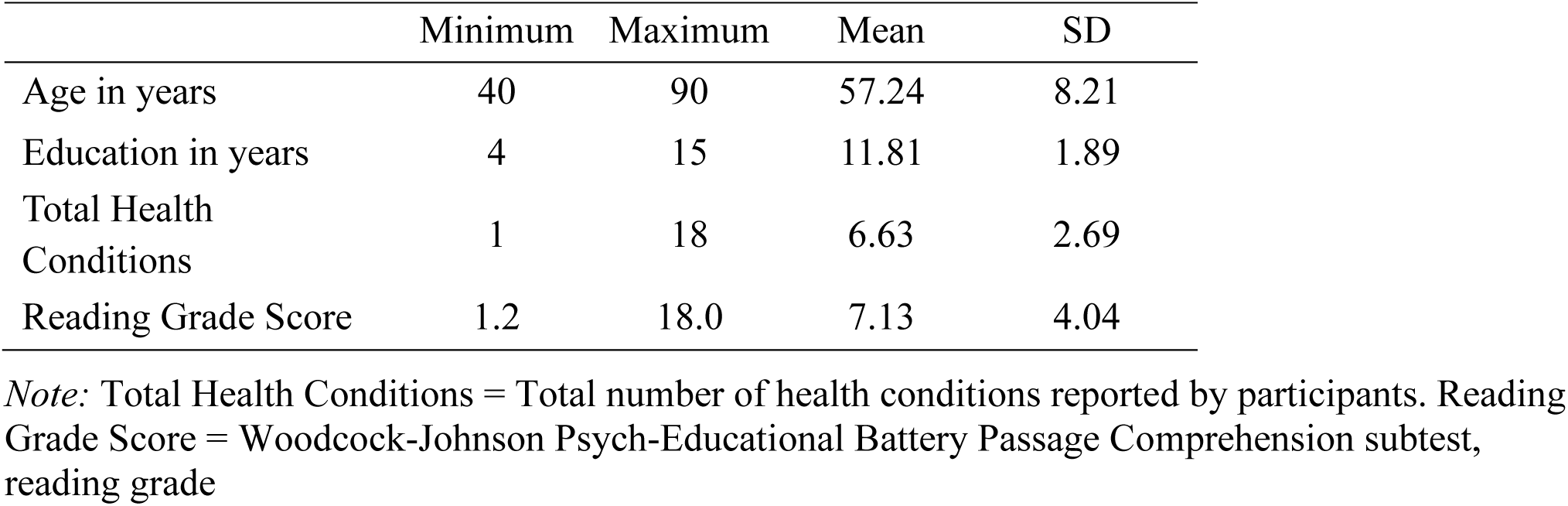
Descriptive statistics for the sample (n = 275)

### 3.2 Item Characteristics

Individual item statistics are presented in Table 3. Item-level descriptive statistics (Table 2) indicated uniformly high mean scores, ranging from 4.19 to 4.32 on a 5-point scale. All items showed moderate to strong negative skew (−1.23 to −1.62) and positive kurtosis (3.09–4.90), suggesting that participants tended to endorse the upper end of the response scale and that responses were tightly clustered. This pattern reflects generally favorable perceptions of information tailoring and limited variability among respondents. This pattern is consistent with the scale’s intent and the intervention’s design, but may constrain the ability to detect associations with other variables due to reduced variability.

**Table 3.** Fit indices for single factor and bifactor confirmatory analyses.

|  | CFI | TLI | RMSEA | SRMR |
| --- | --- | --- | --- | --- |
| One Factor | 0.92 | 0.89 | 0.17 | 0.05 |
| Bifactor | 0.96 | 0.93 | 0.14 | 0.04 |

*Note:* CFI = confirmatory fit index; TLI = Tucker-Lewis index; RMSEA = root mean square error of approximation; SRMR = standardized root mean square residual

Inter-item correlations are presented in Table 4. Correlations among the eight SIT items were all positive, statistically significant (all *p*s < 0.01), and of moderate to strong magnitude (Table 3). Correlations ranged from *r* = 0.51 to 0.83, with a median of approximately .70, indicating substantial coherence among items. The strongest associations were observed between items reflecting the applicability and thought-provoking qualities of the information (“I could use the information” and “Gave me something to think about,” *r* = 0.83). In contrast, the weakest associations involved relational or engagement-oriented items (e.g., “I could relate to the information” and “I want to try out some of the things,” *r* = 0.51). This pattern suggests that while the items assess different aspects of perceived tailoring, they share a core consistent with a single underlying construct.

**Table 4.** Inter-item correlations.

|  | I could<br>relate to the<br>information | About<br>people<br>like me | Easy to<br>understand | I could use<br>the<br>information | Gave me<br>something<br>to think<br>about | About<br>problems<br>people like<br>me have | Would<br>work for<br>me | I want to<br>try out |
| --- | --- | --- | --- | --- | --- | --- | --- | --- |
| I could relate to the<br>information | 1.00 | 0.69 | 0.70 | 0.54 | 0.53 | 0.57 | 0.57 | 0.51 |
| About people like me | 0.69 | 1.00 | 0.74 | 0.75 | 0.73 | 0.72 | 0.62 | 0.56 |
| Easy to understand | 0.70 | 0.74 | 1.00 | 0.75 | 0.74 | 0.68 | 0.68 | 0.64 |
| I could use the<br>information | 0.54 | 0.75 | 0.75 | 1.00 | 0.83 | 0.78 | 0.75 | 0.66 |
| Gave me something<br>to think about | 0.53 | 0.73 | 0.74 | 0.83 | 1.00 | 0.81 | 0.75 | 0.70 |
| About problems<br>people like me have | 0.57 | 0.72 | 0.68 | 0.78 | 0.81 | 1.00 | 0.75 | 0.68 |
| Would work for me | 0.57 | 0.62 | 0.68 | 0.75 | 0.75 | 0.75 | 1.00 | 0.78 |
| I want to try out<br>some of the things | 0.51 | 0.56 | 0.64 | 0.66 | 0.70 | 0.68 | 0.78 | 1.00 |

*Note:* All correlations are significantly different from zero (all $ps < 0.01$ ).

### 3.3 Exploratory Factor Analysis (EFA)

Exploratory analyses with parallel analysis suggested a single-factor solution (Figure 1). Most SIT items loaded well on the first factor. Table 5 lists the factor loadings for the one-factor solution. Overall, the pattern of loadings is consistent with a unidimensional structure.

**Figure 1.**
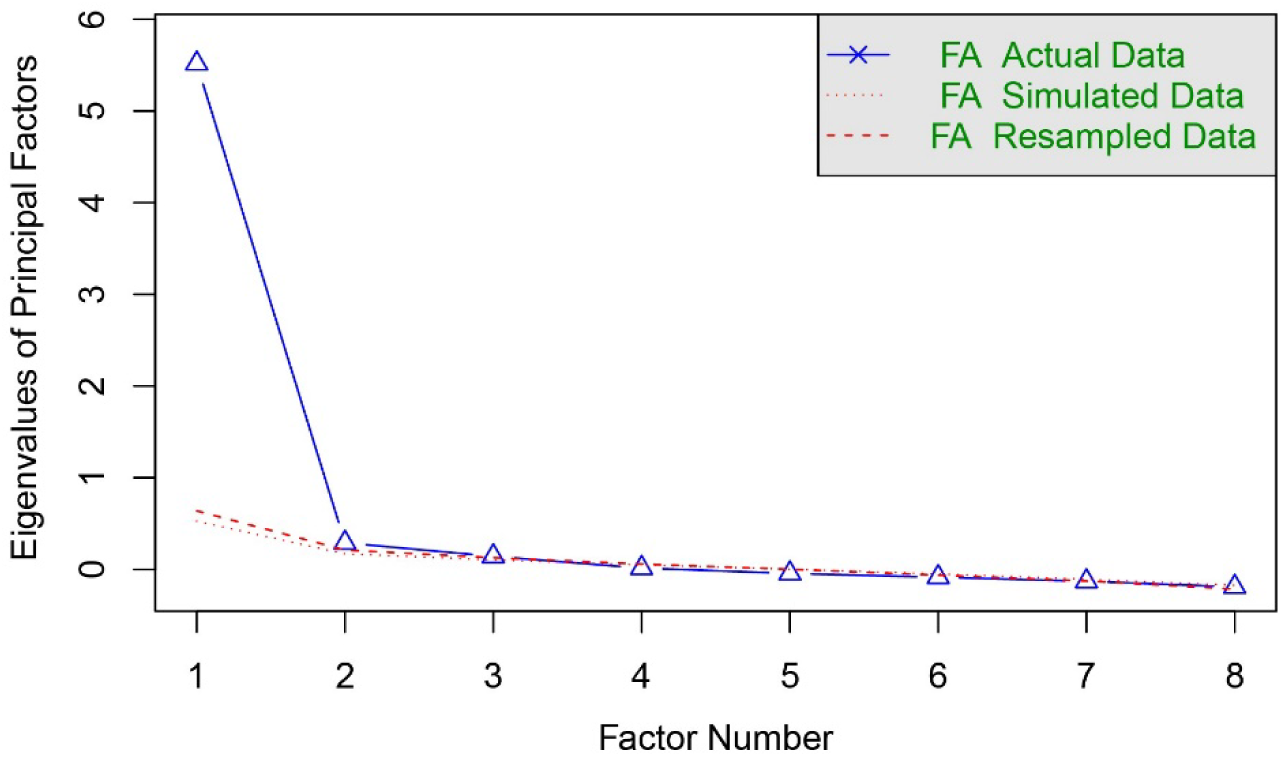
Scree plot of parallel factor analysis *Note*: FA = Factor analysis

**Figure 2.**
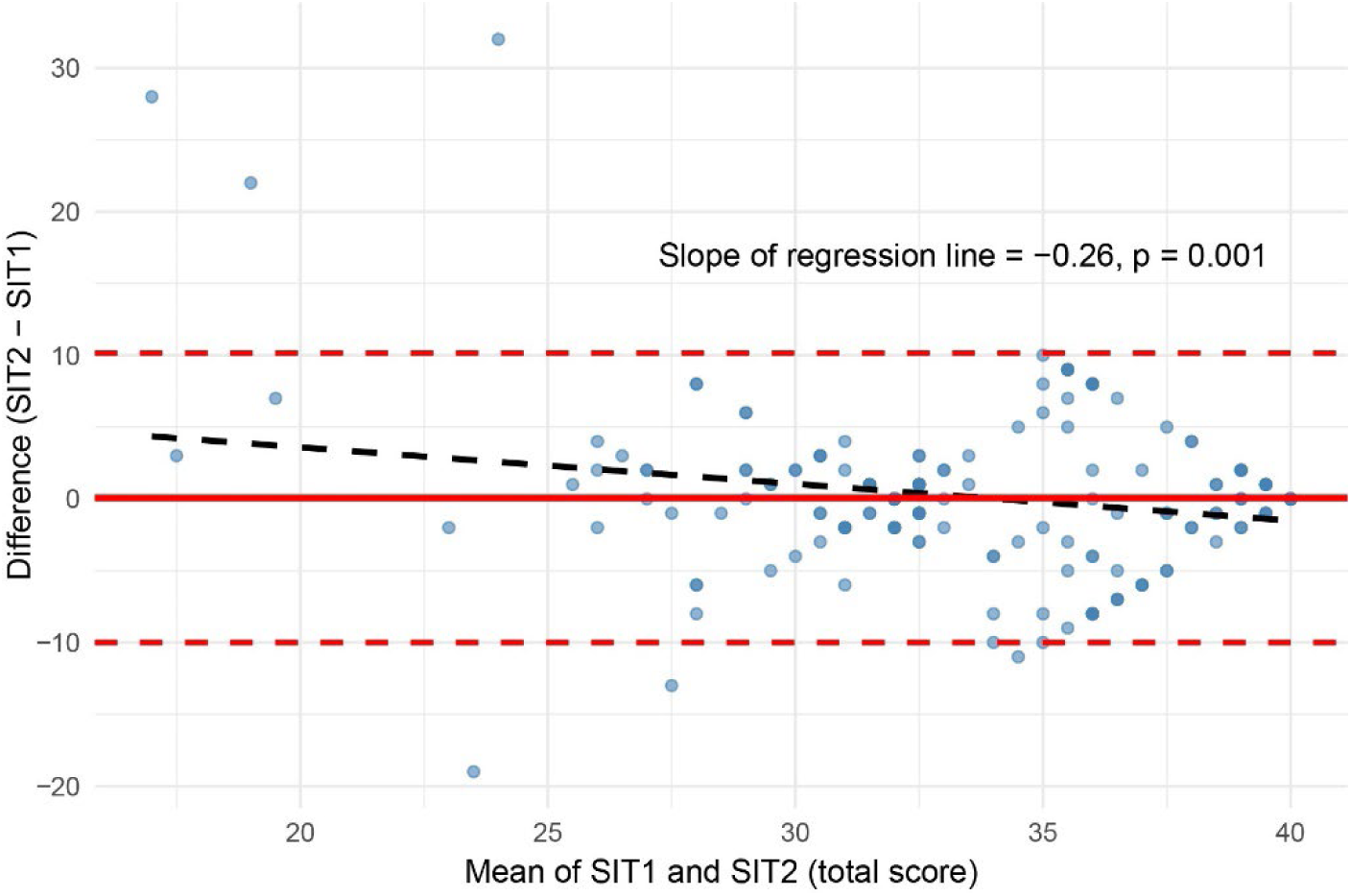
Bland–Altman plot with regression line.

**Table 5.** Exploratory factor loadings.

|  | Loading |
| --- | --- |
| I could relate to the information | 0.67 |
| About people like me | 0.82 |
| Easy to understand | 0.83 |
| I could use the information | 0.90 |
| Gave me something to think about | 0.90 |
| About problems people like me have | 0.87 |
| Would work for me | 0.84 |
| I want to try out some of the things | 0.77 |

### 3.4 Confirmatory Factor Analysis (CFA)

To further assess the factor structure of the SIT, we fit a single-factor confirmatory factor analytic model. It fit the data moderately well (CFI = 0.92, TLI = 0.89, RMSEA = 0.17, SRMR = 0.05) but not at an acceptable level. Considering this evidence of misfit, we explored several alternate factor models; the one that fit the data best was a bifactor model with one general and two minor factors. The first minor factor included the items “The information would work for me” and “I want to try out some of the things talked about,” while the second included “The information was easy to understand” and “I could relate to the information.” Fit indices were substantially improved (CFI = 0.96, TLI = 0.93, RMSEA = 0.14, SRMR = 0.04). The bifactor model fit the data significantly better than the single-factor model (likelihood ratio test χ^2^ [4] = 42.22, *p* < 0.001).

The bifactor model thus provided the best overall fit among competing structures. Model-level indices indicated that variance in SIT responses was dominated by a strong general factor representing perceived tailoring. The general factor accounted for 88% of the common variance (ECV = 0.88), with a high proportion of uncontaminated correlations (PUC = 0.93). Reliability coefficients were excellent (ΩT = 0.96; ΩH = 0.93), indicating that most reliable variance in total SIT scores reflected the general factor. Construct replicability (H = 0.96) and factor determinacy (FD = 0.98) were also high, supporting stable estimation of the general factor. In contrast, the specific factors showed low unique reliability (ΩHS = 0.18–0.22) and modest replicability (H = 0.28–0.38), suggesting limited distinct variance beyond the general factor. Together, these results indicate that the SIT functions mainly as a unidimensional measure of perceived tailoring, and that a single total score provides a psychometrically useful representation of the construct.

### 3.5 Longitudinal Measurement Invariance

Configural, metric, and scalar models were compared for the bifactor model across administrations (Table 6). Fit indices changed only slightly moving from configural to metric invariance and stayed within the cutoffs. The CFI and RMSEA changed minimally from configural to metric models (Δ CFI = −.01, Δ RMSEA = 0.00; Δχ² [8] = 8.31, *p* = 0.40), indicating equivalent factor loadings across assessments. These results supported metric invariance, suggesting that the SIT items operated in a consistent way over time with similar factor loadings. However, fit indices did not support scalar invariance. Constraining item intercepts (scalar model) reduced fit substantially (Δ CFI = −.05, Δ RMSEA = +.02), consistent with the significant χ² difference (Δχ² [6] = 131.16, *p* < 0.001). Scalar invariance was thus not supported.

**Table 6.**
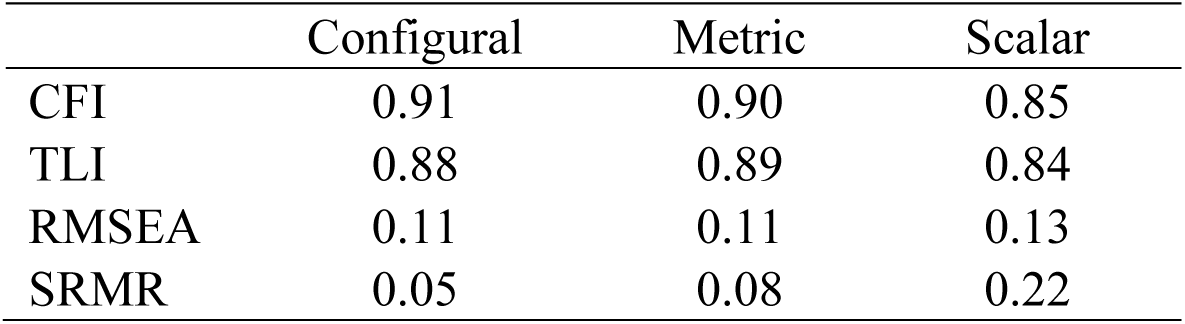
Fit indices for factorial invariance models.

### 3.6 Reliability Analyses

Test–retest reliability across administrations was moderate. The intraclass correlation coefficient (ICC) was calculated as a single measure with a two-way random-effects in both absolute-agreement and consistency models. The ICC for absolute agreement [ICC(A,1)] was 0.60 (95% CI 0.51-0.69), and the consistency ICC [ICC(C,1)] showed a similar pattern (ICC = 0.61, 95% CI 0.52-0.70). The standard error of measurement (SEM) was 3.63, giving a smallest detectable change (SDC) of 10.07. Cronbach’s alpha for the scale at time 1 was 0.95; at time 2, it was 0.93. McDonald’s omega was 0.95. at time 1 and 0.93 at time 2.

A Bland–Altman plot was used to examine agreement between the two SIT administrations. The mean difference between scores was close to zero. However, regressing score differences on their means revealed a statistically significant negative slope (β = −0.26, *p* = .001), suggesting a proportional bias: participants with lower average SIT scores tended to show slightly higher second-administration scores, whereas those with higher average scores showed smaller or slightly negative differences. Despite this trend, most differences fell within the 95% limits of agreement, indicating good overall test–retest consistency. A spaghetti plot of individual SIT scores (Figure 3) shows variability across participants in the two administrations, even though the group means remained stable (red line in Figure 3).

**Figure 3.**
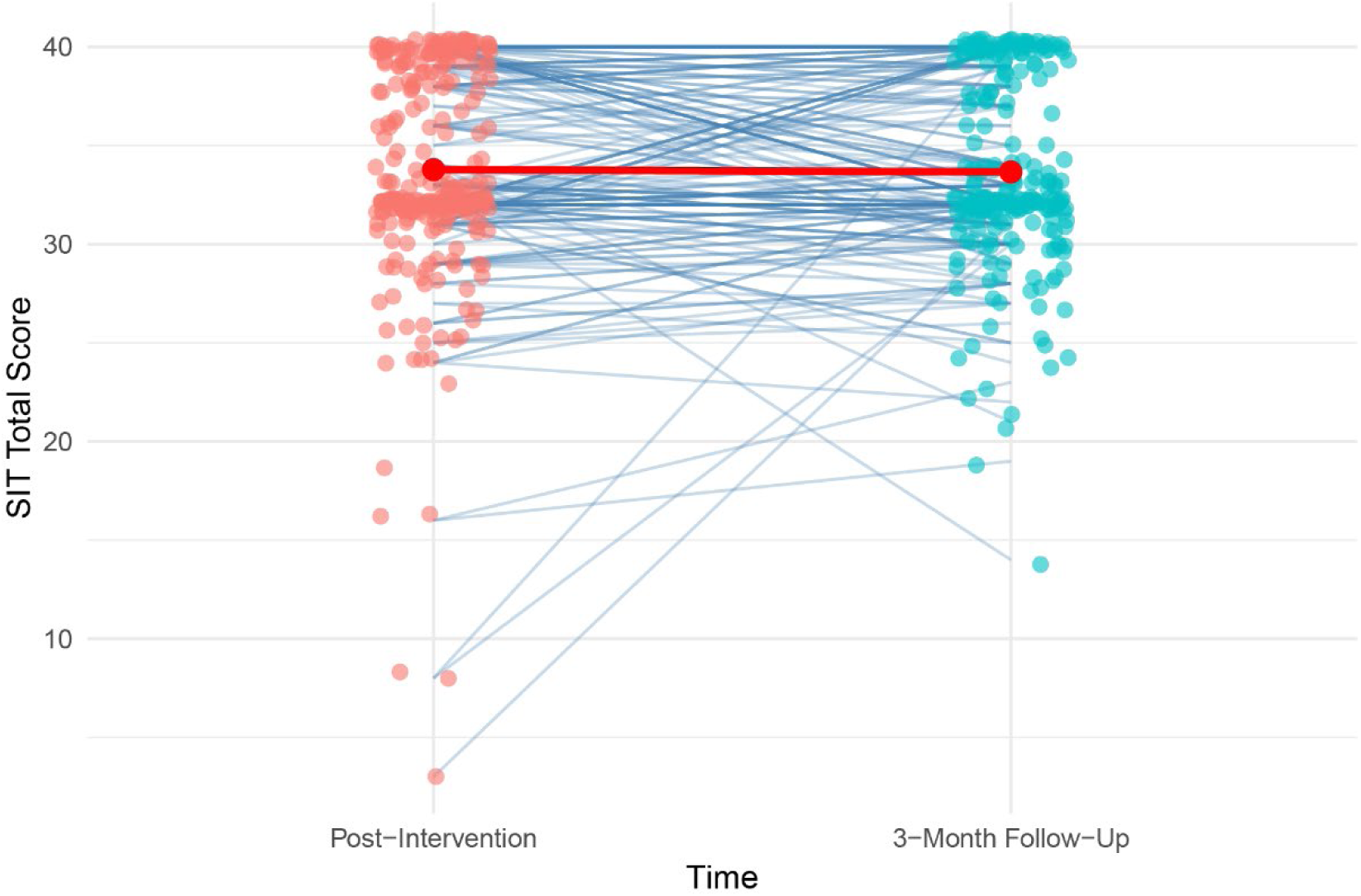
Spaghetti plot of SIT1 vs. SIT2 scores

### 3.7 Convergent and Divergent Validity

As expected, SIT scores correlated positively with measures assessing related constructs (Figure 4. Perceived tailoring was most strongly associated with participants’ ratings of the information’s usefulness (User Useful; *r* = .33, *p* < .001) and with patient activation (PAM; *r* = .27, *p* < .001).

**Figure 4.**
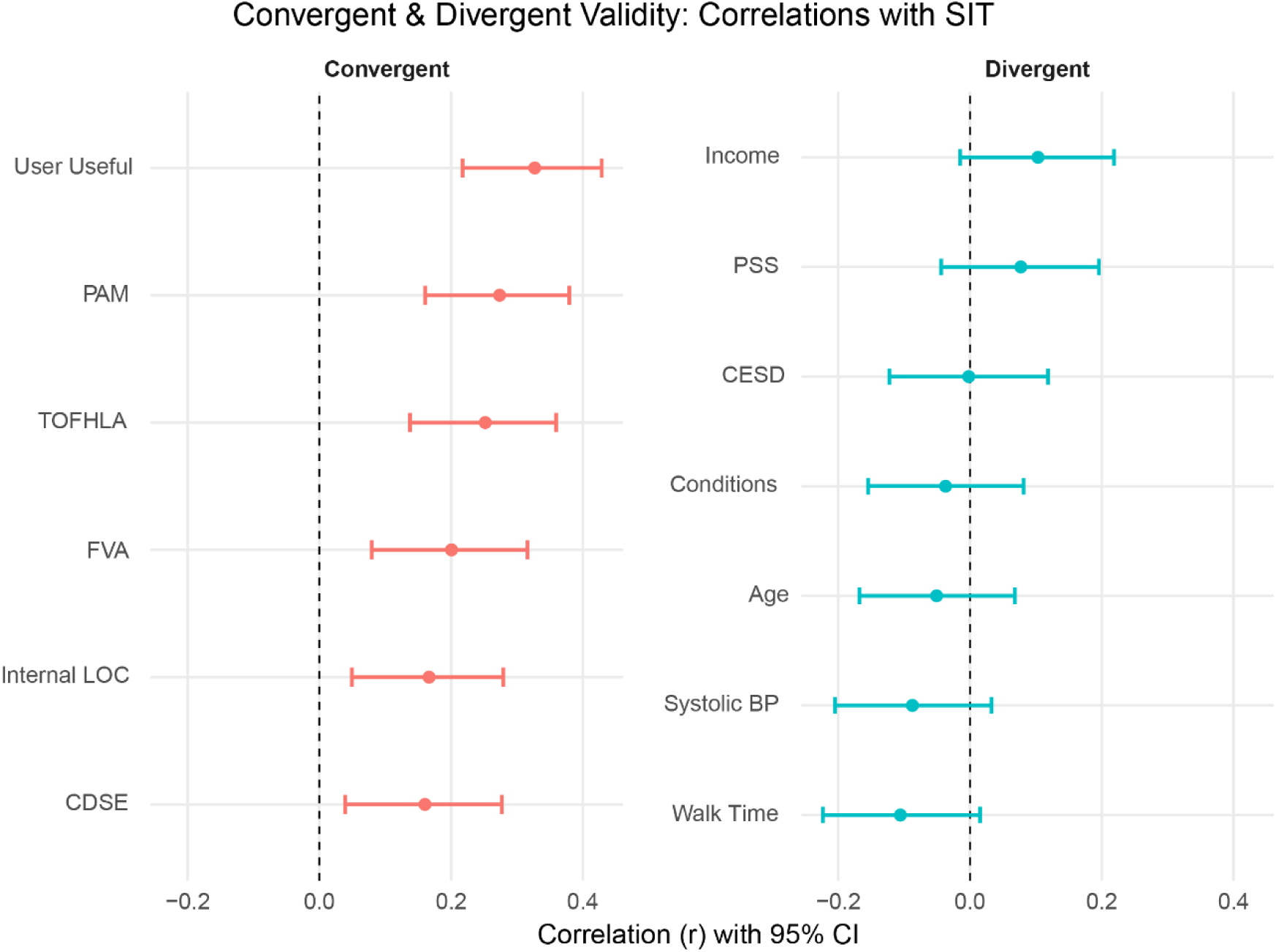
Correlations of SIT with convergent and divergent measures. *Note:* User Useful = Usefulness scale of Technology Acceptance Model scale; PAM = Patient Activation Measure; TOFHLA = Test of Functional Health Literacy in Adults; FVA = FLIGHT/VIDAS Health Literacy Measure; Internal LOC = Internal control subscale of the Multidimensional Health Locus of Control Scale; CDSE = Chronic Disease Self-Efficacy Scale; Income = Participant self-report of income; PSS = Perceived Stress Scale; CESD = Center for Epidemiological Studies—Depression scale; Conditions = Participant report of total number of chronic health conditions; Age = age in years; Systolic BP = Participant systolic blood pressure before completing the 10-meter walk test; Walk Time = Time taken in the 10-meter walk test

Smaller but still significant correlations were observed with health literacy (TOFHLA; *r* = .25, *p* < .001; FVA; *r* = .20, *p* = .001), internal health locus of control (*r* = .17, *p* = .006), and chronic disease self-efficacy (*r* = .16, *p* = .010). In contrast, SIT scores were not significantly related to demographic and health variables—including income, age, number of chronic conditions, and systolic blood pressure—and to psychological measures of perceived stress (PSS; *r* = .08, ns) and depressive symptoms (CESD; *r* = 0.002, ns). This pattern demonstrates good convergent validity with conceptually related psychosocial constructs and divergent validity with unrelated health and demographic factors.

### 3.8 Predictive Validity

The latent growth models incorporating patient activation (PAM) and chronic disease self-efficacy (CDSE) showed that SIT scores obtained immediately after the intervention predicted change in outcomes over time. Figure 5 shows the growth curve for patient activation. Patient activation changed significantly over the course of the study (coefficient = 0.17, SE = 0.08, *z* = 2.08, *p* = 0.04). The regression of the model’s slope on the SIT score was also significant (coefficient = 0.05, SE = 0.02, *z* = 3.32, *p* = 0.001) as was the regression of the QOL measure (SF36 General Health scale at three-month follow-up (coefficient = 14.73, SE = 5.51, *z* = 2.67, *p* = 0.007). The indirect effect of the SIT on QOL via change in activation was also statistically significant (estimate = 0.71, SE = 0.23, *z* = 3.16, *p* = 0.002).

**Figure 5.**
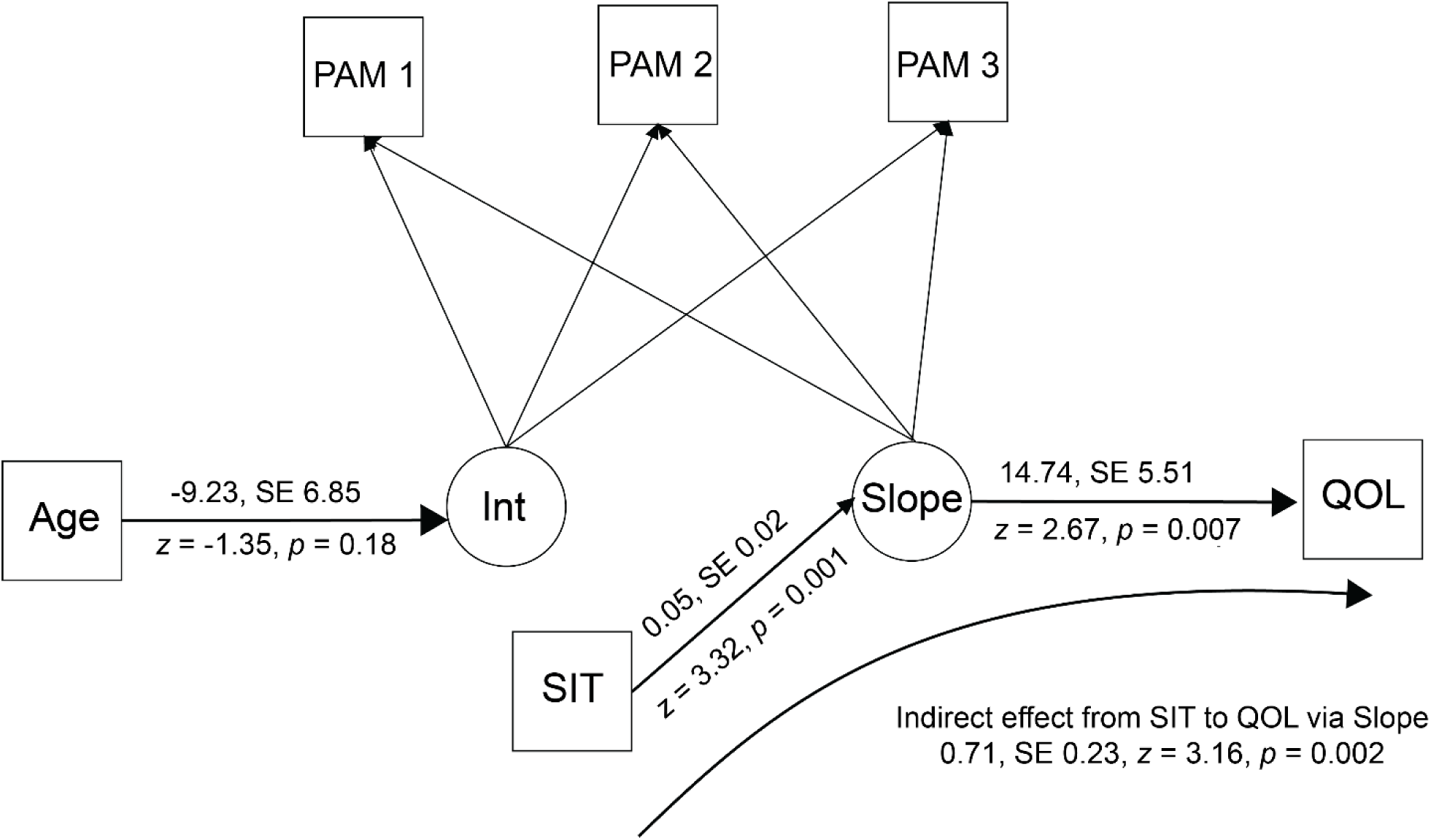
Latent growth curve model for patient activation. *Note:* PAM = Patient Activation Measure; Age = age in years; Int = model intercept; Slope = model slope; QOL = SF36 General Health scale at three-month follow-up; SIT = Success in Tailoring scale score

Figure 6 shows the growth curve for chronic disease self-efficacy. It increased significantly over the course of the study (coefficient = 1.26, SE = 0.27, *z* = 4.64, *p* < 0.001). The regression of the model’s slope on the SIT score was also significant (coefficient = 0.13, SE = 0.06, *z* = 2.26, *p* = 0.02), but the regression of the QOL measure (SF36 General Health scale at three-month follow-up; (coefficient = −0.11, SE = 0.08, *z* = −1.42, *p* = 0.15) was not. The indirect effect of the SIT on QOL was not statistically significant (estimate = 0.05, SE = 0.04, *z* = 1.26, *p* = 0.21).

**Figure 6.**
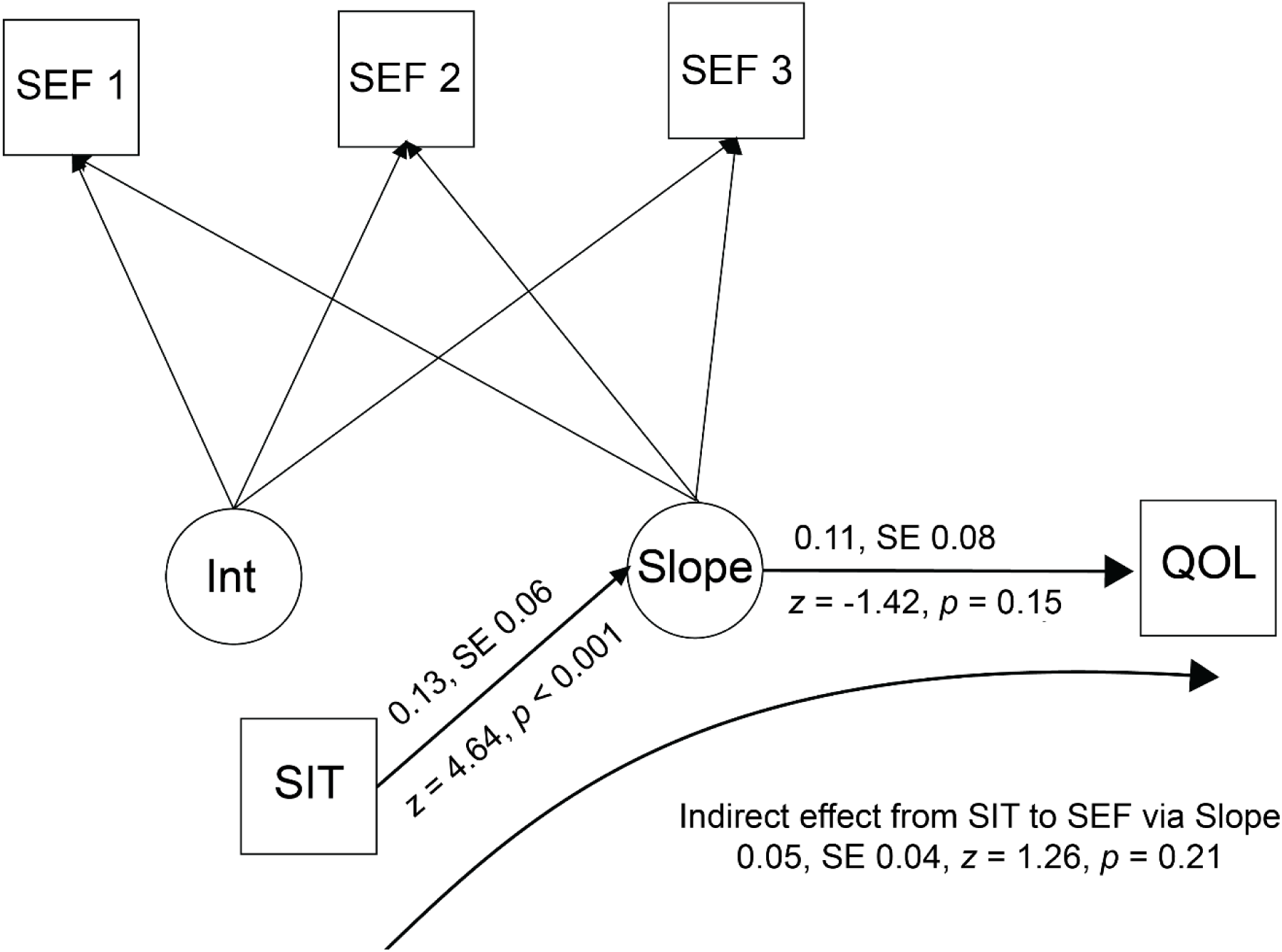
Latent growth curve model for self-efficacy. *Note:* SEF = Chronic Disease Self-Efficacy Scale; Int = model intercept; Slope = model slope; QOL = SF36 General Health scale at three-month follow-up; SIT = Success in Tailoring scale score

## 4 Discussion

### 4.1 Overview

The purpose of this paper was to provide more detailed information on the SIT’s reliability and validity to help support its potential usefulness. We conclude that the SIT does what it was designed to do: it provides a method to measure users’ impressions of tailored health information in apps.

Factor analyses were consistent with a single common factor, reliability is adequate, and the validity analyses are consistent with our hypotheses. SIT scores obtained immediately after the intervention were related to the pattern of change in the key outcomes of patient activation and chronic disease self-efficacy.

### 4.2 Factor Structure

Although the bifactor model yielded the best statistical fit, the pattern of indices clearly supports the practical use of a single SIT composite score. The strong general factor accounted for a large portion of the common variance, while the specific factors contributed little unique and reliable information. This suggests that participants’ responses primarily reflect an overarching perception that the information they received was well tailored to their needs, rather than distinct dimensions of tailoring. Accordingly, the SIT can be interpreted and scored as a unidimensional measure of perceived tailoring, simplifying its use in both research and applied settings while retaining conceptual coherence with the elaboration likelihood model’s emphasis on personal relevance as the key mechanism of tailored communication.

### 4.3 Measurement Invariance and Reliability

Test–retest reliability, measured by ICCs, was moderate. The Bland–Altman plot showed a small but significant average bias but individual variability. In practical terms, the group average was stable, although changes in a single person’s score must be interpreted with care. SEM and SDC estimates provide useful guidance on interpreting changes in SIT scores.

### 4.4 Validity

The correlations of measured to represent similar and unrelated constructs were consistent with hypotheses. The SIT correlated significantly with participants’ ratings of the app’s usefulness and with the PAM and CDSE. These findings are consistent with the hypothesized relation as the underlying commonality is that both are about how people see their ability to act on health information. Correlations with unrelated measures were close to zero, supporting divergent validity.

### 4.5 Predictive Validity

The latent growth curve models showed that people with higher baseline SIT scores made greater changes in activation and disease management self-efficacy over the course of the study. The SIT did not predict initial levels (intercepts); it only predicted the slope of improvement. This suggests that feeling the information was relevant and usable was associated with greater change later.

### 4.6 Limitations

There are a few caveats. The sample was persons 40 years and older with chronic illness and low health literacy, so results may not apply to other groups. Follow-up was only three months. CFA fit indices were not ideal, and ICCs were moderate, so the scale is better suited for group-level research than individual monitoring. Missing data also may have influenced some results.

The negatively skewed distributions of SIT items indicate that participants overwhelmingly perceived the intervention content as well tailored to their needs. While such skewness and kurtosis are expected in user-centered interventions, they may also signal ceiling effects that constrain variability and attenuate correlations with other constructs. Future refinements to the SIT might explore expanding the response range or incorporating items that better differentiate among individuals with uniformly positive perceptions. Nevertheless, the pattern of high endorsement supports the scale’s face validity and aligns with the intervention’s design to deliver highly personalized information.

Although the bifactor analysis indicated strong general-factor dominance, some multidimensionality may still exist, and future research should examine whether distinct facets of perceived tailoring emerge in different populations or intervention contexts.

### 4.7 Implications and Future Directions

Even with these limitations, the SIT gives researchers and program designers a simple way to determine whether efforts to tailor health information to users are successful. It provides a means of assessing mechanisms of behavior change in digital health interventions. Future studies should examine how the scale performs in other populations, assess longer-term predictive validity, and explore the smaller factors suggested by the CFA results.

## 6 Author Contributions

RLO: Conceptualization, data curation, formal analysis, funding acquisition, investigation, project administration, supervision, writing – original draft; RD: Data curation, investigation, project administration, resources, supervision, writing – revise and edit; JC: Conceptualization, investigation, writing – revise and edit

## Data Availability

All data produced in the present study are available upon reasonable request to the authors.

The raw data supporting the conclusions of this article will be made available by the authors on request, without undue reservation.

